# Intraoperative glucocorticoids confound surgical biopsy transcriptomics

**DOI:** 10.64898/2026.09.08.26362140

**Authors:** Frederik Tibert Larsen, Charlotte Wilhelmina Wernberg, Jyotsna Nambiar, Subhajit Dutta, Elise Jonasson, Lea Ladegaard Grønkjær, Birgitte Gade Jacobsen, Susanne Mandrup, Anne Loft, Søren Fisker Schmidt, Mette Munk Lauridsen, Kim Ravnskjaer, Lars Grøntved

**Author notes:** Equal contribution, order determined by mutual agreement (Frederik Tibert Larsen, Charlotte Wilhelmina Wernberg). The authors have declared that no conflict of interest exists.

## Abstract

Human tissue transcriptomic studies frequently rely on biopsies obtained during surgery under general anesthesia, yet the impact of perioperative medications on gene expression has not been systematically evaluated. Using paired liver biopsies from patients with obesity at risk for metabolic dysfunction–associated steatohepatitis (MASH), we compared RNA sequencing profiles from biopsies collected under local anesthesia months before surgery with matched intraoperative biopsies obtained under general anesthesia. Intraoperative samples exhibited striking transcriptional changes enriched for canonical glucocorticoid receptor target genes, implicating perioperative dexamethasone administration as the principal driver. In a matched cohort in which dexamethasone was administered immediately after biopsy collection, we directly demonstrate that dexamethasone profoundly alters hepatic gene expression during surgery. These findings were independently validated and shown to substantially distort associations between liver fibrosis stage and inflammatory gene expression, obscuring biologically relevant immune signatures. Similar glucocorticoid-responsive transcriptional changes were observed in adipose tissue, indicating a systemic effect. Our findings identify perioperative dexamethasone exposure as a previously unrecognized but readily modifiable confounder in human tissue transcriptomics. As biopsy-based bulk and single-cell transcriptomic studies continue to expand, careful documentation and consideration of perioperative medication exposure will be essential to improve the reproducibility, interpretability, and biological validity of human gene expression studies.

## Introduction

Transcriptomic analyses of biopsies have provided unprecedented insights into cell state transitions across diverse diseases(1), including metabolic dysfunction-associated steatotic liver disease (MASLD)(2). Many biopsies are obtained intraoperatively under general anesthesia, where glucocorticoids, such as dexamethasone (DEX), are routinely administered perioperatively to prevent postoperative nausea and vomiting (PONV). (3). As a potent glucocorticoid receptor agonist, DEX rapidly alters transcription of hundreds of metabolic and inflammatory genes within 0.5–2 hours(4, 5), meaning its intraoperative use can profoundly bias transcriptomic analyses.

## Results and discussion

To assess whether intraoperative use of DEX influence biopsy transcriptomes, we performed RNA-seq on liver biopsies from donors at risk of MASLD undergoing weight-loss surgery. This included a paired preoperative percutaneous biopsy (non-surgical) and an intraoperative laparoscopic biopsy (surgical) collected months apart (Figure 1A). All intraoperative procedures followed the same anesthesia protocol; except that one group received DEX only after biopsy collection (termed W/O DEX), whereas the other received DEX at anesthesia induction (termed DEX). The groups did not differ in baseline characteristics and time between biopsies (Suppl. Table 1). This paired study design enabled assessment of the intraoperative effects of DEX while adhering to ethical considerations.

**Figure. 1:**
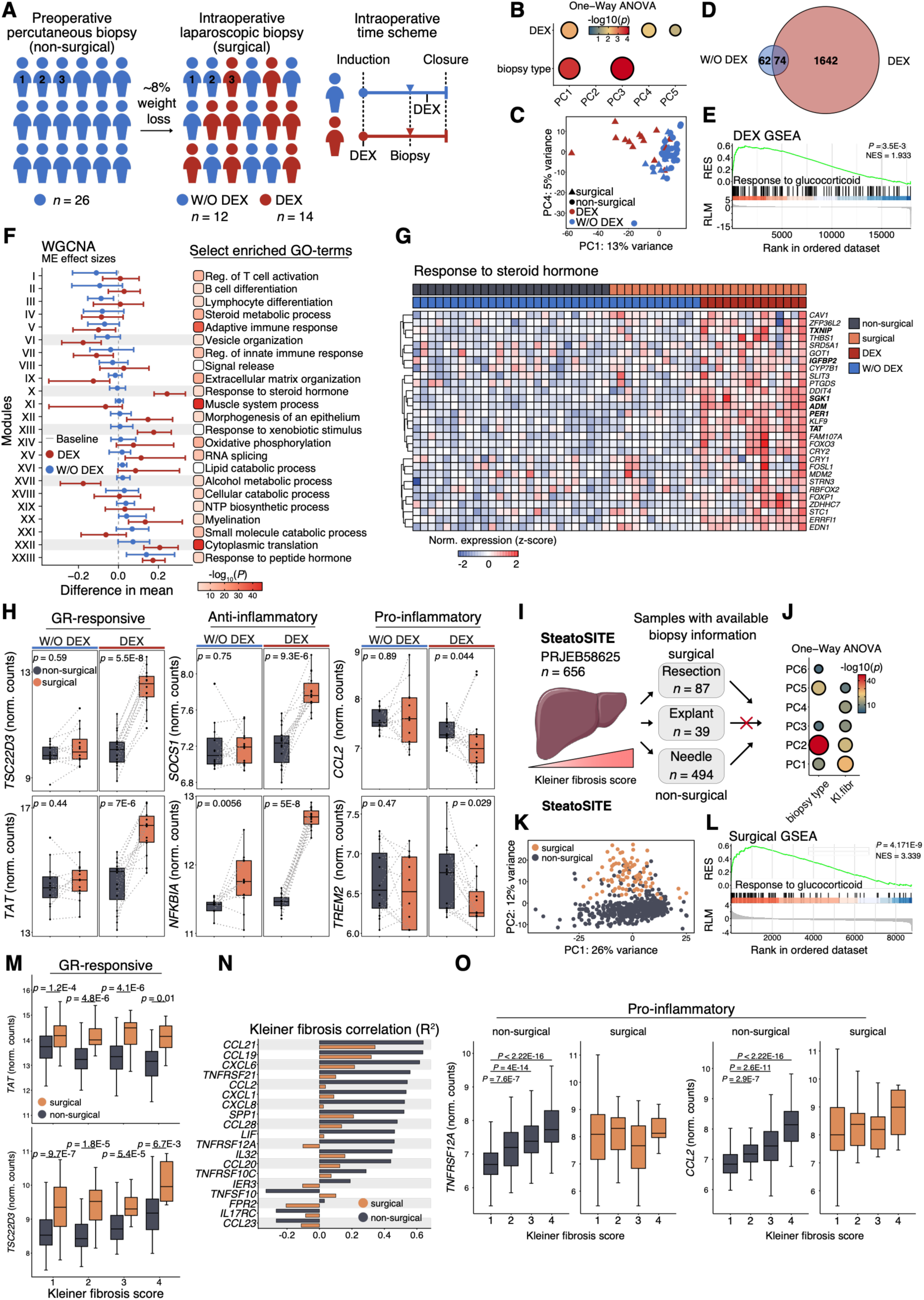
Impact of perioperative DEX on hepatic transcriptomes. (**A**) Study cohort enrolled for weight-loss surgery. (**B**) Effect of non-surgical versus surgical biopsy sampling and intraoperative DEX treatment. P-value is shown in colorbar as -log10(p). (**C**) PCA comparing expression profiles by intraoperative DEX treatment. (**D**) Overlap of DEGs between paired non-surgical and surgical biopsies with and without intraoperative DEX treatment. (**E**) GSEA using DEGs between paired non-surgical and surgical biopsies. (**F**) Difference in mean effect sizes between paired non-surgical and surgical biopsies of WGCNA-identified module eigengenes with representative enriched GO terms shown. (**G**) Hierarchical clustering of module X genes associated with fat cell differentiation. (**H**) Expression of representative glucocorticoid-responsive, anti-inflammatory, and pro-inflammatory genes. (**I**) Available SteatoSITE RNA-seq data. (**J**) Effect of biopsy type and fibrosis stage. (**K**) PCA comparing expression profiles by biopsy type. (**L**) Top GSEA result using DEGs between resection and needle biopsies. (**M**) Expression of representative glucocorticoid-responsive genes across Kleiner fibrosis grades grouped by biopsy type. (**N**) Pearson correlation coefficients of top pro- and anti-inflammatory genes correlating with Kleiner fibrosis grade grouped by biopsy type. (**O**) Representative proinflammatory genes across Kleiner fibrosis grades. Mann-Whitney U test and Wilcoxon signed-rank tested difference in distribution between non-paired and paired groups, respectively.

Principal component analysis (PCA) of transcriptomes revealed marked transcriptional differences between biopsy collection procedures, with clear DEX-associated effects on PC1, PC3, and PC5 (Figure 1B–C). For paired non-surgical versus surgical biopsies, this included 1,716 differentially expressed genes (DEGs) with DEX and 136 DEGs in W/O DEX (Figure 1D, Suppl. Fig. 1A, Suppl. Data A-B). This DEX effect was independent of statistical thresholds (Suppl. Fig. 1B). Consistently, gene set enrichment analysis (GSEA) revealed significant enrichment of glucocorticoid response-related genes in surgical biopsies from DEX donors (Figure 1E, Suppl. Data C) which could not be adjusted for by modelling neither by covariate adjustment nor by empirical bayes corrected counts (Suppl. Fig. 1C). For genes regulated independently of DEX, we found enrichment of inflammatory pathways in agreement with moderate weight loss between biopsy collection (Suppl. Fig. 1D and Suppl. Data D).

To investigate the biological impact of DEX treatment, we applied weighted gene co-expression network analysis (WGCNA). GO-enrichment analysis showed enrichment of biological pathways in 20 of 23 of the identified modules (Figure 1F). Calculated effect sizes of module eigengenes between paired non-surgical and surgical biopsies revealed significant DEX regulation of pathways involved in steroid signaling response, cytoplasmic translation, vesicle organization, and alcohol metabolic process (Figure 1F, Suppl. Data E). Genes associated with steroid signaling response were induced in donors given DEX intraoperatively (Figure 1G). Specifically, surgical DEX-treated donors showed induction of classical GR-responsive genes such as *TSC22D3* and *TAT* and anti-inflammatory GR-responsive genes such as *SOC1* and *NFKBIA*, and repression of proinflammatory genes such as *CCL2* and *CCL8* (Figure 1H). Importantly, transcriptomics analysis of adipose tissue biopsies from the same cohort revealed similar significant effects of DEX, suggesting systemic effects (Suppl. Figure 1E-J, Suppl. Data F-H).

Together, these findings reveal profound transcriptomic differences between non-surgical biopsies and surgical biopsies collected in the presence of DEX. Notably, most recent liver biopsy transcriptomics studies that uses surgical samples do not report glucocorticoid use, nor anesthesia protocols (Suppl. Table 2I). To address this, we contacted several anesthesiologists at hospitals across Europe and the US. Seven responded and all reported routine use of glucocorticoids (Suppl. Figure 2) aligned with current guidelines(3).

To test whether transcriptomic data from non-surgical versus surgical biopsies affect correlations between gene expression and liver fibrosis score, we analyzed data from the SteatoSITE cohort(6) (Figure 1I-O and Suppl. Data J-K). Consistent with the severity of MASLD, we observed that Kleiner fibrosis grade influenced PC1-2 (Figure 1J-K). Interestingly, and in agreement with our study, we found that biopsy procedure influenced PC2 and PC5 (Figure 1J-K) and GSEA of DEGs (non-surgical vs. surgical, FDR<0.05, Log2FC 0.1, Suppl. Data J) showed enrichment of glucocorticoid regulated pathways (Figure 1L-M, Suppl. Data K), implying DEX usage as a potential cofounder. Moreover, we found a blunted correlation between inflammatory cytokines, chemokines, and receptors and Kleiner fibrosis grades in surgical biopsies (Figure 1N) and the known incremental increase of *TNFRSF12A* and *CCL2* with Kleiner fibrosis grade was absent in surgical liver biopsies (Figure 1O).

Collectively, these findings show that perioperative glucocorticoids influence transcriptomic profiles in surgical biopsies and may represent an important confounder. We advocate for transparent reporting of perioperative glucocorticoid use and, whenever feasible, administration after biopsy collection. These considerations are likely relevant for transcriptomic studies of any tissue obtained under general anesthesia.

## Data Availability

All data produced in the present study are available upon reasonable request to the authors

## Methods

Details on data analysis are described in supplemental material.

## Supplemental material

### Supplemental data

- Suppl. Data: Transcriptomic data analysis (separate excel file)

## Supplemental figures and tables

**Suppl. Fig. 1:**
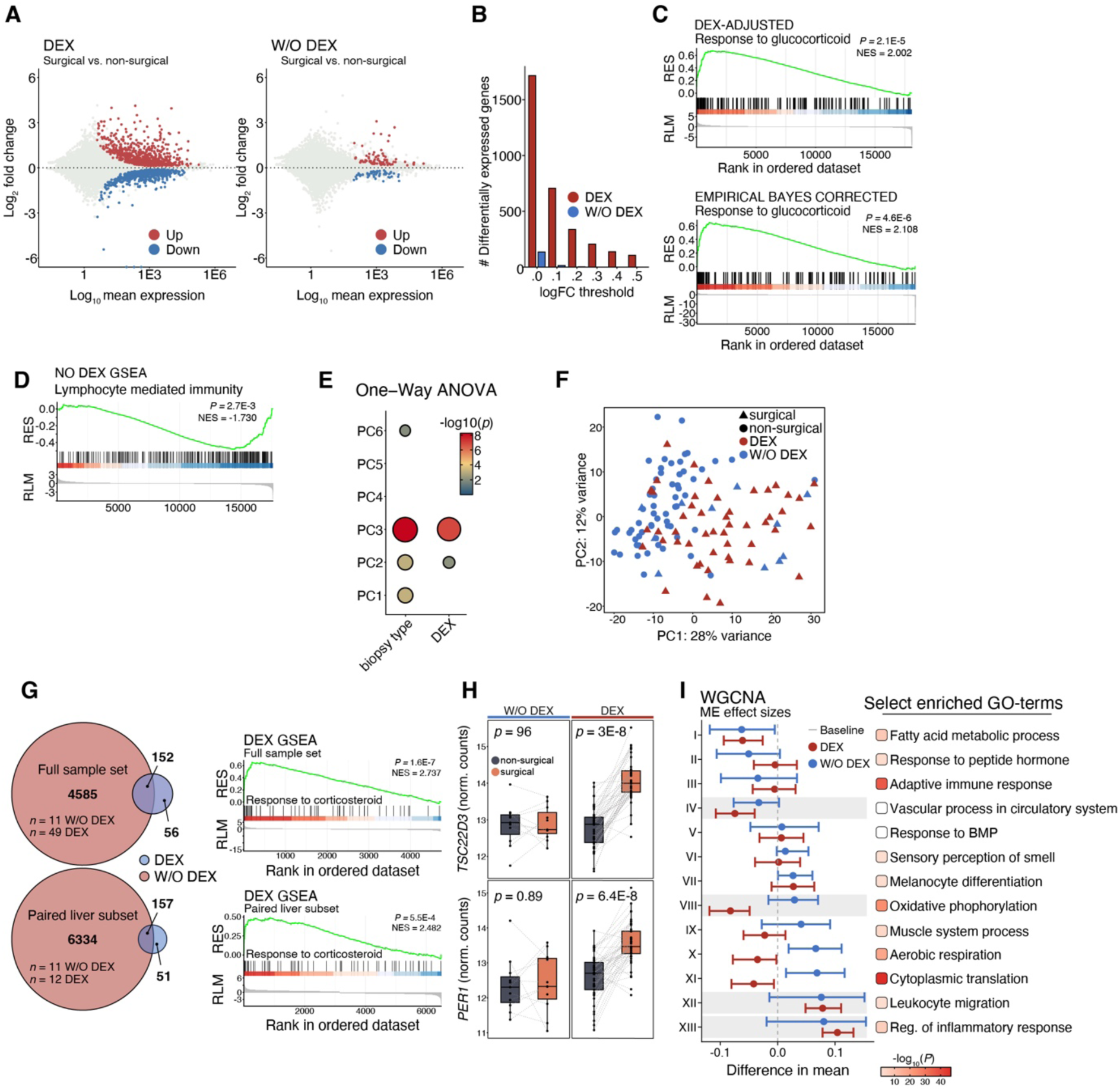
Impact of perioperative DEX on liver (panel A-D) and adipose tissue (panel E-I) transcriptomes. **(A)** MA-plot showing log2 fold changes of differentially expressed genes color in red and blue. **(B)** Number of differentially expressed genes detected with increasing effect sizes (lfcThreshold 0 – 0.5). **(C)** Top results from gene set enrichment analysis (GSEA) using results from differential genes expression analysis adjusted for DEX exposure (upper) or from differential gene expression analysis on empirical bayes corrected counts (lower). **(D)** GSEA using DEG result between paired non-surgical and surgical biopsies without intraoperative DEX treatment. **(E)** Effect of non-surgical versus surgical biopsy sampling and intraoperative DEX treatment. P-value is shown in colorbar as -log10(p). **(F)** Principal component analysis (PCA) comparing expression profiles by intraoperative DEX treatment. **(G)** Overlap of differentially expressed genes (DEGs) between paired non-surgical and surgical biopsies with and without intraoperative DEX treatment for full sample set (n = 11 w/o DEX, n = 49 DEX) and patients paired to liver biopsies used (n = 11 w/o DEX, n = 12 DEX) with top results from GSEA using DEGs between paired non-surgical and surgical biopsies for the full sample set (n = 4737 genes) and patients paired to liver biopsies used (n = 6491 genes). **(H)** Expression of representative glucocorticoid-responsive genes. Wilcoxon signed-rank test was employed to test difference in distribution between paired groups. **(I)** Difference in mean effect sizes between paired non-surgical and surgical biopsies of WGCNA-identified module eigengenes with representative enriched GO terms shown. Top 5 modules are highlighted.

**Suppl. Figure. 2:**
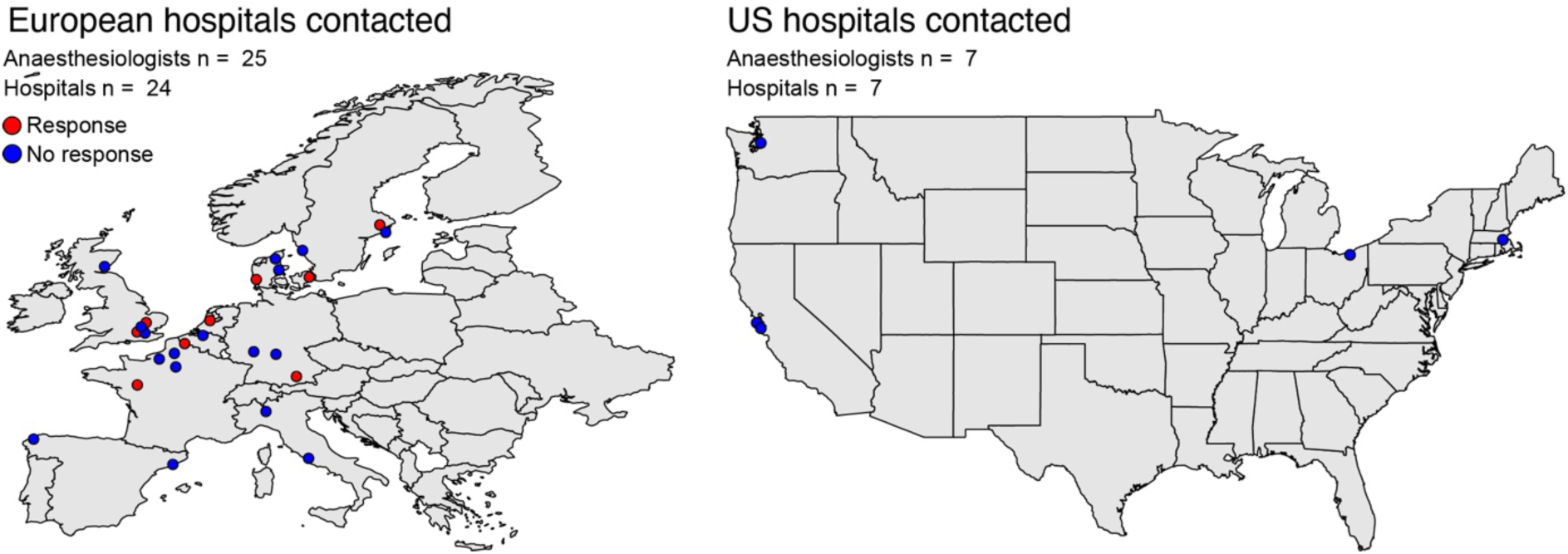
Geographical distribution of hospitals in Europe and the United States with affiliated anesthesiologists who were invited to participate in the survey. Lead and consulting anesthesiologists at hospitals reported to have contributed samples to transcriptomic studies included in the reviewed manuscript were contacted, and to broaden the scope, additional anesthesiologists at other hospitals in Europe and the United States were also approached. In total, the survey was distributed via email to 33 anesthesiologists across 12 countries (France, England (UK), Scotland (UK), the United States, Germany, Belgium, the Netherlands, Italy, Spain, Denmark, and Sweden). The questions asked were: 1. Is dexamethasone routinely used for PONV prophylaxis? 2. Is it primarily administered for antiemetic, anti-inflammatory, analgesic, or other purposes? 3. For which type of surgery is it typically used? 4. Does your department perform bariatric surgery? 5. At what point in the perioperative period is it usually administered (e.g., pre-induction, intraoperatively, postoperatively)? Red and blue dots represent hospitals from which a response or no response, respectively, was received from affiliated anesthesiologists.

**Suppl. Table 1.** Biometric and biochemistry variables in the patient cohort in mean ± standard deviation.

|  | DEX |  | W/O DEX |  |
| --- | --- | --- | --- | --- |
|  | non-surgical | surgical | non-surgical | surgical |
| n | 14 | 14 | 12 | 12 |
| Sex (M/F) | 4/10 | 4/10 | 2/10 | 2/10 |
| Age (years) | 41.6±10.0 | 42.0±10.7 | 43.9±10.1 | 44.2±10.8 |
| BMI (kg/m<sup>2</sup>) | 43.2±5.6 | 40.8±5.4 | 43.4±5.0 | 40.5±4.2 |
| ALT (U/L) | 49.4±43.9 | 36.6± 22.6 | 40.3±33.8 | 37.6± 22.6 |
| AST (U/L) | 29.6±8.4 | 31.2±10.5 | 32.7±22.8 | 29.9±10.9 |
| HDL | 1.1±0.6 | 1.0±0.1 | 1.1±0.2 | 1.3±0.2 |
| LDL | 3.2±1.1 | 3.0±0.9 | 2.6±0.8 | 3.1±0.9 |
| Cholesterol (mmol/L) | 4.7±1.6 | 4.4±1.1 | 4.0±0.7 | 4.6±0.7 |
| Triglycerides (mmol/L) | 1.8±1.0 | 1.6±0.7 | 1.4±0.5 | 1.2±0.5 |
| FIB-4 | 0.8±0.5 | 0.9±0.6 | 0.7±0.3 | 0.7±0.3 |
| C-peptide (pmol/L) | 1,425.6±407.4 | 1,251.4±262.8 | 1,185.1±594.0 | 1,223.7±710.9 |
| HOMA-IR | 7.3±3.5 | 4.9±1.9 | 5.5±4.2 | 6.7±7.6 |
| LSM (kPa) | 8.5±3.9 | 7.6±4.1 | 6.4±3.2 | 7.0±3.1 |
| CRP (mg/mL) | 5.9±6.0 | 7.9±10.1 | 7.9±6.5 | 9.1±6.6 |

WLS, weight-loss surgery. ALT, alanine aminotransferase. AST, aspartate aminotransferase. APRI, AST-to-platelet ratio index. CRP, C-reactive protein. FIB-4, fibrosis-4. HDL, high density lipoprotein. HOMA-IR, homeostatic model assessment for insulin resistance. LDL, low density lipoprotein. LSM, liver stiffness measurement.

**Suppl. Table 2.**
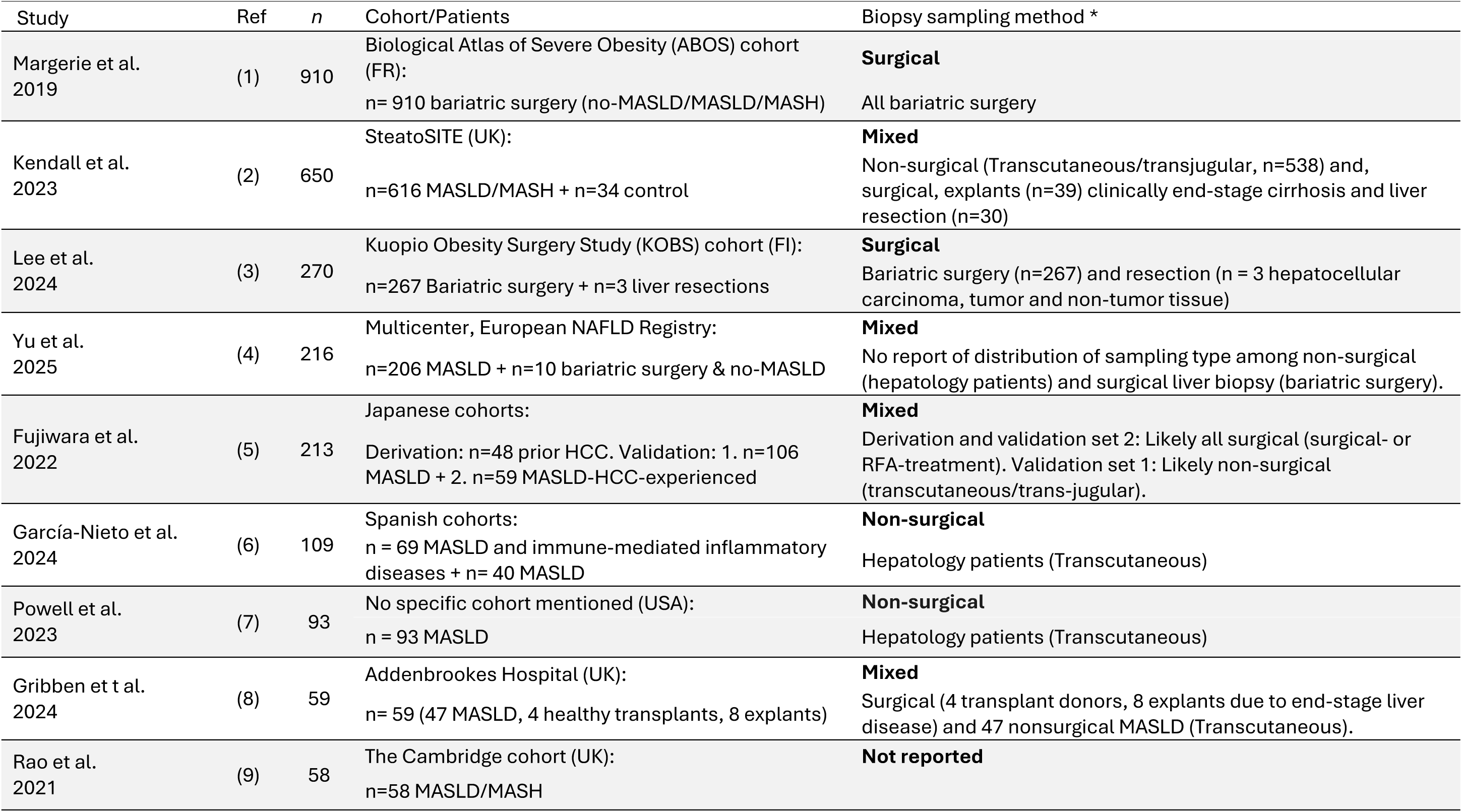
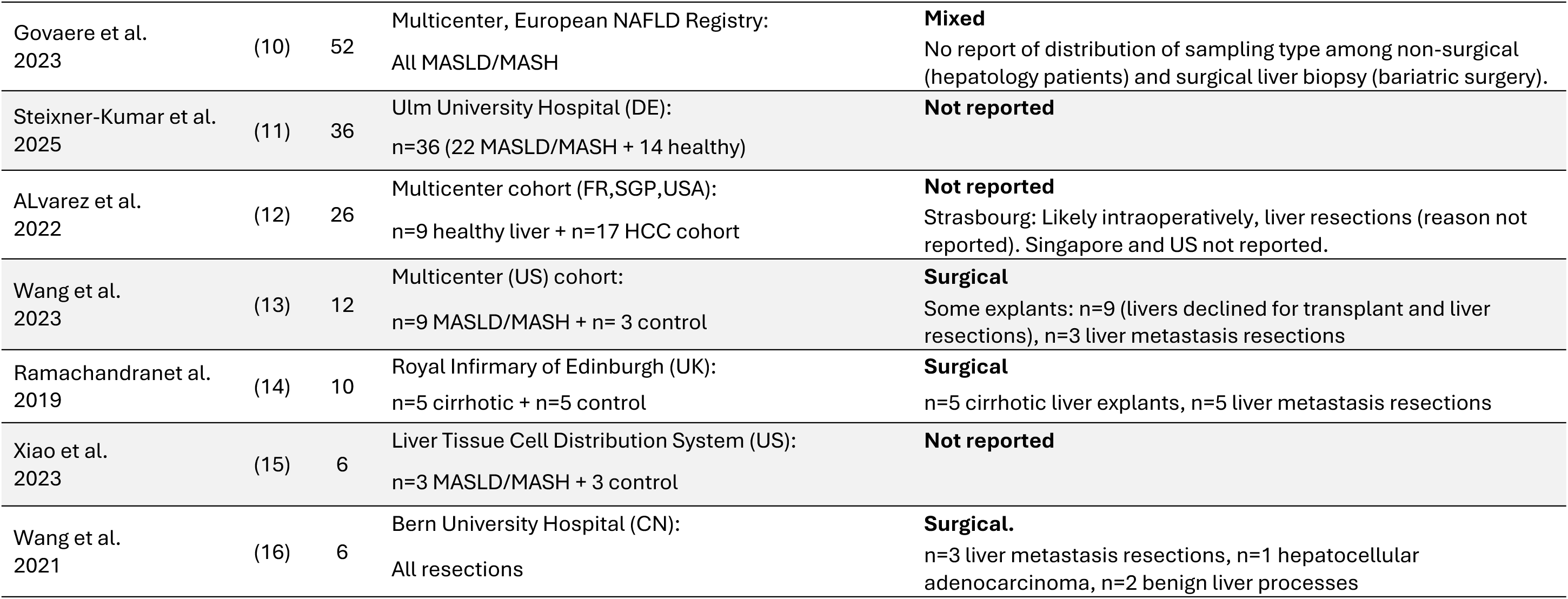
Studies on human liver transcriptome and information on the type of biopsy. *Information on methods of tissue sampling was extracted through supplementary material of articles, which is almost never available in the main text. RFA, Radiofrequency ablation.

| Study | Ref | n | Cohort/Patients | Biopsy sampling method * |
| --- | --- | --- | --- | --- |
| Margerie et al. 2019 | (1) | 910 | Biological Atlas of Severe Obesity (ABOS) cohort (FR):<br>n= 910 bariatric surgery (no-MASLD/MASLD/MASH) | **Surgical**<br>All bariatric surgery |
| Kendall et al. 2023 | (2) | 650 | SteatoSITE (UK):<br>n=616 MASLD/MASH + n=34 control | **Mixed**<br>Non-surgical (Transcutaneous/transjugular, n=538) and, surgical, explants (n=39) clinically end-stage cirrhosis and liver resection (n=30) |
| Lee et al. 2024 | (3) | 270 | Kuopio Obesity Surgery Study (KOBS) cohort (FI):<br>n=267 Bariatric surgery + n=3 liver resections | **Surgical**<br>Bariatric surgery (n=267) and resection (n = 3 hepatocellular carcinoma, tumor and non-tumor tissue) |
| Yu et al. 2025 | (4) | 216 | Multicenter, European NAFLD Registry:<br>n=206 MASLD + n=10 bariatric surgery & no-MASLD | **Mixed**<br>No report of distribution of sampling type among non-surgical (hepatology patients) and surgical liver biopsy (bariatric surgery). |
| Fujiwara et al. 2022 | (5) | 213 | Japanese cohorts:<br>Derivation: n=48 prior HCC. Validation: 1. n=106 MASLD + 2. n=59 MASLD-HCC-experienced | **Mixed**<br>Derivation and validation set 2: Likely all surgical (surgical- or RFA-treatment). Validation set 1: Likely non-surgical (transcutaneous/trans-jugular). |
| García-Nieto et al. 2024 | (6) | 109 | Spanish cohorts:<br>n = 69 MASLD and immune-mediated inflammatory diseases + n= 40 MASLD | **Non-surgical**<br>Hepatology patients (Transcutaneous) |
| Powell et al. 2023 | (7) | 93 | No specific cohort mentioned (USA):<br>n = 93 MASLD | **Non-surgical**<br>Hepatology patients (Transcutaneous) |
| Gribben et t al. 2024 | (8) | 59 | Addenbrookes Hospital (UK):<br>n= 59 (47 MASLD, 4 healthy transplants, 8 explants) | **Mixed**<br>Surgical (4 transplant donors, 8 explants due to end-stage liver disease) and 47 nonsurgical MASLD (Transcutaneous). |
| Rao et al. 2021 | (9) | 58 | The Cambridge cohort (UK):<br>n=58 MASLD/MASH | **Not reported** |
| Govaere et al.<br>2023 | (10) | 52 | Multicenter, European NAFLD Registry:<br>All MASLD/MASH | **Mixed**<br>No report of distribution of sampling type among non-surgical (hepatology patients) and surgical liver biopsy (bariatric surgery). |
| Steixner-Kumar et al.<br>2025 | (11) | 36 | Ulm University Hospital (DE):<br>n=36 (22 MASLD/MASH + 14 healthy) | **Not reported** |
| ALvarez et al.<br>2022 | (12) | 26 | Multicenter cohort (FR,SGP,USA):<br>n=9 healthy liver + n=17 HCC cohort | **Not reported**<br>Strasbourg: Likely intraoperatively, liver resections (reason not reported). Singapore and US not reported. |
| Wang et al.<br>2023 | (13) | 12 | Multicenter (US) cohort:<br>n=9 MASLD/MASH + n= 3 control | **Surgical**<br>Some explants: n=9 (livers declined for transplant and liver resections), n=3 liver metastasis resections |
| Ramachandran et al.<br>2019 | (14) | 10 | Royal Infirmary of Edinburgh (UK):<br>n=5 cirrhotic + n=5 control | **Surgical**<br>n=5 cirrhotic liver explants, n=5 liver metastasis resections |
| Xiao et al.<br>2023 | (15) | 6 | Liver Tissue Cell Distribution System (US):<br>n=3 MASLD/MASH + 3 control | **Not reported** |
| Wang et al.<br>2021 | (16) | 6 | Bern University Hospital (CN):<br>All resections | **Surgical.**<br>n=3 liver metastasis resections, n=1 hepatocellular adenocarcinoma, n=2 benign liver processes |

## Supplemental methods

### Sex as a biologic variable

The study includes male and female donors of liver and adipose tissue biopsies. Sex of the doners was not considered for the paired longitudinal transcriptomic analysis.

### Study design and participants

This study is an exploratory RNAseq analysis of liver biopsies and adipose tissue to describe the transcriptomic response to dexamethasone in patients undergoing bariatric surgery. It included participants with risk factors of MASLD and serial liver biopsy from a prospective case–control study in patients with severe obesity (BMI ≥35 kg/m2), previously described in detail(17). Patients included in this study all underwent bariatric surgery (Roux-en-Y gastric bypass or sleeve gastrectomy). Baseline visits preceded surgery, with a mean of 346 days (SD = 197 days). During this period the patients lost 5-8% of their body weight required for enrolment to bariatric surgery. No alternative liver diseases were suspected or found, and participants were included based on risk factors, without prior non-invasive liver assessment. Following ethical approval, all participants were offered a liver biopsy at baseline and at the surgery visit. Recruitment and study visits were conducted at the University Hospital of Southern Denmark, Esbjerg (2018– 2025). Participants were at least 18 years old and provided written informed consent. Exclusion criteria: other liver diseases, self-reported alcohol consumption >20 g/day/week (women) or >30 g/day/week (men), decompensated cirrhosis, use of hepatotoxic medications, pregnancy, or malignant diseases. All investigations adhered to a standardized protocol during both visits. This included liver biopsy, medical history, transient elastography (TE) using FibroScan (Echosens, Paris, France), and blood sampling, all of which were conducted on the same day under fasting conditions. The trials complied with GCP standards and the Declaration of Helsinki. Ethical approval was obtained from the regional ethics authorities (S-20160006 G). Data management utilized Redcap via the Open Patient Data Explorative Network (Odense, DK)

### Tissue and blood sampling

All measurements and samples were collected the same day as liver biopsies. Liver stiffness measurement scans were performed by experienced staff using FibroScan (Echosens, Paris, France). Experienced staff used M-/XL probes as indicated. A reliable measurement was defined as having at least 10 valid measurements and an IQR of <30% if the LSM was >7.1 kPa. Blood was drawn by trained staff, and all routine biochemical analyses were performed by local and central laboratories using commercially available kits.

### Liver and adipose tissue biopsies

Baseline percutaneous biopsies from the right liver lobe were obtained using a 16–18G suction needle. During weight-loss surgery, intraoperative biopsies were collected with an 18G automatic biopsy gun (True-Core). DEX was administered prior to biopsy retrieval (induction) in 14 patients as part of standard PONV prophylaxis, while 12 patients received DEX after the surgical liver and adipose tissue biopsy were collected.

### Histology and staging of MASLD

All liver biopsies were analyzed by one expert hepatopathologist (TC), who was blinded to all other data. Histology was graded using NAS (0–8) Clinical Research Network: steatosis (0–3), lobular inflammation (0–3), and ballooning (0–2). Fibrosis staging used the Kleiner fibrosis score (0–4). Biopsies were considered sufficient if ≥10 mm in length with six or more portal tracts or regenerative nodules.

### RNA-sequencing and preprocessing

Liver needle biopsies and subcutaneous adipose tissue biopsies preserved in RNA-later were homogenized using the FastPrep-24™ system (MP Biomedicals, Irvine, CA). RNA was extracted with TRIzol RNA lysis reagent (#T9424, Thermo Fisher Scientific, Waltham, MA) according to the manufacturer’s protocol. RNA concentration was measured using the Qubit 3.0 Fluorometer (Thermo Fisher Scientific), and RNA integrity was assessed with a Fragment Analyzer 5200 (Agilent, Santa Clara, CA). Library preparation was performed using the NEBNext Ultra RNA Library Prep Kit for Illumina (New England Biolabs, San Diego, CA), following the manufacturer’s instructions. RNA was paired-end sequenced using the NovaSeq™ 6000 platform (Illumina, San Diego, CA). Sequencing reads were aligned to the human reference genome (GRCh38, Ensembl release 110) using STAR (v2.7.8a)(18). Exon-level read quantification was carried out with FeatureCounts (v2.0)(19). Quality control of sequencing data was performed using FastQC and MultiQC. To remove genetic variants from the raw sequencing reads, BAMboozle (v0.5.0) was applied(20). Sanitized reads have been deposited in the NCBI Gene Expression Omnibus repository and are accessible through accession number GSE320593.

### Data analysis of RNA-sequencing data

Batch effect was corrected for using a negative binominal model implemented in ComBat-seq from the sva package (v.3.56)(21). Corrected count data was used for differential gene expression analysis using DESeq2 (v.1.48.2)(22). Differentially expressed genes were identified using a Wald test and a-error accumulation from multiple testing were adjusted for using Benjamini-Hochberg correction implemented in the package. Adjusted p values (p.adj.) ≤ 0.05 were considered statistically significant. Gene counts were normalized for further analysis using variance stabilized transformation (vst). Modules of co-expressed genes were identified using the weighted gene co-expression network analysis (WGCNA) package (v.1.70)(23). Briefly, vst-normalized counts (n = 10000 transcripts) with top loadings across the first four principal components (PCs) were chosen for analysis. By using scale free topology analysis, a soft threshold power of 12 was chosen for calculating the topological overlap matrix. Dynamic tree cut method was set to hybrid with a deep split of 2 and a minimum cluster size of 20 genes. Estimation statistics were employed to calculate effects sizes with 95% confidences intervals for module eigenes between paired baseline and weight-loss surgery donor samples were calculated using the dabestr package(24). Biological functions of gene sets and modules were explored using gene set-enrichment analysis and gene ontology (GO) enrichment analysis implemented in the R package clusterProfiler (v.4.16.0)(25).

For the transcriptomic analysis of liver biopsies from the SteatoSITE cohort(2) we excluded explant samples to avoid a confounding effect from the severe underlying liver disease and stratified samples into percutaneous needle biopsies (non-surgical, n = 494) and intraoperative resection biopsies (surgical, n = 87).

### Review of existing studies analyzing liver transcriptome

We reviewed studies published between 2019 and 2025 on transcriptomic analysis of liver tissue. Studies were identified through a targeted search of the PubMed database We applied the following filters: English, humans, adult: 19+ years, and excluded preprints. The aim was to evaluate sampling method of the biopsy (i.e., non-surgical, and surgical) and to investigate whether the biopsies were obtained under conditions that could potentially influence the liver transcriptome (such as sedation or drug administration during intraoperative procedures). Studies included a variety of liver conditions, such as metabolic dysfunction associated steatotic liver disease (MASLD), metabolic dysfunction associated steatohepatitis (MASH), hepatocellular carcinoma (HCC) as well as control populations. We extracted data regarding the number and type of patients (and name of cohort if mentioned), method of tissue sampling, and if the type of anesthesia used were described.

### Survey to Anesthesiologic departments, use of dexamethasone

Following completion of the review, we tried to contact lead and consulting anesthesiologists at the hospitals reported to have contributed samples to the transcriptomic studies. To expand the scope of the survey, we additionally approached anesthesiologists at other hospitals in Europe and the US. In total, the survey was distributed via email to 32 anesthesiologists across ten countries (France, the United Kingdom, the United States, Germany, Belgium, the Netherlands, Italy, Spain, Denmark, and Sweden). The questions asked were: 1. Is dexamethasone routinely used for PONV prophylaxis? 2. Is it primarily administered for antiemetic, anti-inflammatory, analgesic, or other purposes? 3. For which type of surgery is it typically used? 4. Does your department perform bariatric surgery? 5. At what point in the perioperative period is it usually administered (e.g., pre-induction, intraoperatively, postoperatively)?

### Statistical analysis

The Shapiro-Wilk test was employed to test for normality. For normal-distributed data, the students t-test was employed. The Mann-Whitney U test was used for multiple comparison of normalized RNAseq count data. Bonferroni Hochberg correction was employed to adjust for a-error accumulation. For multiple comparisons, a nominal p-value ≤ 0.05 was considered statistically significant. Estimation statistics was applied to evaluate module eigengene differences in mean between paired non-surgical and surgical liver biopsies (24). All statistical analyses were performed in R (v. 4.5.1).

